# The Impact of Artificial Intelligence on Large Vessel Occlusion Stroke Detection and Management: A Systematic Review Meta-analysis

**DOI:** 10.1101/2024.03.03.24303653

**Authors:** Elan Zebrowitz, Sonali Dadoo, Paige Brabant, Anaz Uddin, Esewi Aifuwa, Danielle Maraia, Mill Etienne, Neriy Yakubov, Myoungmee Babu, Benson Babu

**Author notes:** **Correspondence to:** Benson Babu MD MBA, Department of Hospital Medicine, Wyckoff Medical Center Brooklyn, NY, USA 11237. Equal Contribution (Elan Zebrowitz, Sonali Dadoo, Paige Brabant, Anaz Uddin).

## Abstract

**Introduction:** Stroke remains the second leading cause of death worldwide, with many survivors facing significant disabilities. In acute stroke care, the timeless adage ‘Time is brain’ underscores the vital need for quick action. Innovative Artificial Intelligence (AI) technology potentially offers swift detection and management of acute ischemic strokes, leading acute stroke care towards enhanced automation.

**Methods:** The study is registered with Prospero under CRD42024496716 and adheres to the Problem, Intervention, Comparison, and Outcomes framework (PICO). The analysis used Preferred Reporting Items for Systematic Reviews and Meta-Analyses (PRISMA) guidelines. We searched Embase, PubMed, DBLP, Google Scholar, IEEE Xplore, Cochrane database, IEEE, Web of Science, ArXiv, MedRxiv Web of Science, and Semantic Scholar. The articles included were published between 2019 and 2023. Out of 1,528 articles identified, thirty-eight met the inclusion criteria.

**Results:** We compared AI-augmented Large Vessel Occlusion (LVO) detection and non-AI in various patient processing times related to emergent endovascular therapy in acute ischemic strokes. Triage Time, Door-to-Intervention Notification Time (INR), and Door-to-Arterial Puncture Time revealed an odds ratio (OR) of 0.39 (95% CI: 0.29 - 0.54, p < 0.001), 0.30 30 (95% CI: 0.21 - 0.42, p < 0.001), and 0.50 (95% CI: 0.30 - 0.82, p = 0.007), respectively. CT-to-Puncture-Time and Door-to-CTA-Time yielded an OR of 0.57 (95% CI: 0.31 - 1.04, p = 0.065) and 0.77 (95% CI: 0.37-1.60, p = 0.489), respectively. The Last Known Well (LWK) to Time of Arrival resulted in an OR of 1.15 (95% CI: 0.83 - 1.59, p = 0.409). AI stroke detection sensitivity OR of 0.91 (95% CI: 0.88 - 0.95, p < 0.001), National Institute of Health score (NIHSS) 16.20 (95% CI: 14.96 - 17.45, p = 0.001). Patient Transfer-Times between primary and comprehensive stroke centers generated an OR of 0.98 (95% CI: 0.73 - 1.32, p = 0.893). Similarly, Door-in-Door-Out Time (DIDO) had an OR of 1.19 (95% CI: 0.21 - 6.88, p = 0.848). The results indicated significant differences across several parameters between the AI and non-AI groups.

**Conclusion:** Our findings highlight how AI augments healthcare providers’ ability to detect and manage strokes swiftly and accurately within acute care settings. As these technologies advance and AI becomes more integrated into healthcare systems, longitudinal studies are critical in evaluating its impact on workflow efficiency, cost-effectiveness, and clinical outcomes.

## Introduction 1.0

Stroke continues to rank as the second leading cause of mortality worldwide, with profound disability affecting most survivors. According to a study in 2021, the average hospitalization duration for stroke patients ranged from 11 to 27 days (Rochmah TN, 2021). This prolonged hospital stay contributes to heightened healthcare costs, imposing a considerable financial burden on patients and healthcare systems (Rochmah TN, 2021). Research suggests that stroke-related expenditures per patient can reach $140,048, which includes acute hospitalization, rehabilitation, and long-term care expenses (Gorelick, 2019). Furthermore, Western healthcare systems allocate around 3-4% of their total expenditure toward stroke detection and treatment efforts (Katan M, 2018).

First-time strokes afflict approximately 610,000 individuals annually, with recurrent strokes affecting an additional 185,000 individuals per year (Virani, 2020). The lifetime risk of stroke has escalated over time, underscoring the persistent threat that strokes pose to public health (Virani, 2020) (OMH, 2024).

A recent study revealed a 7.7% increase in stroke-related mortality across all age groups (Virani, 2020). A significant proportion of stroke survivors grapple with reduced mobility, affecting approximately half of all cases, while 26% continue to face residual challenges with activities of daily living (Katan M, 2018). Other co-morbid medical conditions, such as post-stroke depression, afflict approximately one-third of affected individuals (Virani, 2020).

In acute stroke management, the timeless adage “Time is brain “emphasizes the importance of quick action. Integrating Artificial Intelligence (AI) in stroke detection presents an unparalleled opportunity to enhance the current standard of stroke management by optimizing timely interventions. Deep learning algorithms are used to analyze CT head imaging, enabling the identification of patterns indicative of stroke pathology. Furthermore, AI applications encompass various functionalities, including triage management, quantification of infarct volume, surveillance, and prediction of stroke burden (Soun, 2021). Studies have demonstrated significant improvements in stroke detection rates with AI, with computer-aided diagnosis schemes yielding noteworthy performance metrics (Tang, 2011). The use of CAD results in significantly improved sensitivity and specificity (P < 0.005) (Tang, 2011).

Since 2018, FDA-approved AI software programs have been available for the radiologic detection of Large Vessel Occlusion (LVO) (FDA, 2018). Recent research has evaluated the impact of AI on stroke diagnosis and treatment decision-making with promising results. AI-augmented interpretation of cerebral vasculature imaging has shown the potential to enhance patient outcomes while reducing healthcare costs (van Leeuwen, 2021). Statistical models predict substantial yearly cost savings associated with implementing AI in stroke management protocols (van Leeuwen, 2021). This systematic review meta-analysis examines the existing literature to assess the influence of AI on acute stroke care pathways, including acute stroke detection, Triage, patient processing times leading to emergent endovascular therapy, costs, and clinical outcomes.

## Methods 2.0

The study is registered with Prospero under CRD42024496716 and adheres to the PICO framework. Problem: Large Vessel Occlusion (LVO) Detection. Intervention: Artificial Intelligence (AI). Comparison: Performance: AI-augmentation versus non-AI. Outcome:

Door-to-Triage Time, Door-to-CTA Time, CT-to-Puncture Time, Door-to-Arterial Puncture Time, Door-to-Intervention Notification Time (INR), and AI Sensitivity.

The analysis used PRISMA (Preferred Reporting Items for Systematic Reviews and Meta-Analyses) guidelines (MJ, 2020). To identify relevant articles, we comprehensively searched databases, including Embase, PubMed, DBLP, Google Scholar, IEEE Xplore, Cochrane Database, Web of Science, Arxiv, Medrxiv, and Semantic Scholar.

Our search strategy is as follows: 1) Boolean Logic: combined connecting words such as “AND, ““OR, “and “NOT “in various combinations to expand or narrow down search results 2) Fuzzy Logic: search terms such as “AI “NEAR “LVO Detection “or “CT “WITHIN 5 words of “AI “to search for particular articles 3) Truncation: We searched for terms that began with a specific string by placing an asterisk (*) at the end of a root word. For example, “Occlu* “to include “Occluded “and “Occlusion. “The word strings used primary keyword (or phrase) in combination with one or two secondary keywords (for example (((“artificial intelligence”) AND (“Large Vessel Occlusion”)) AND (CTA)); (((“Large Vessel Occlusion”) AND (“AI”) AND (software)) AND (workflow times)); (((“Large Vessel Occlusion”) AND (“AI”) AND (software)) AND ((CTA) AND (door-in))); (((“Large Vessel Occlusion”) AND (“AI”) AND (software)) AND ((accuracy) AND (sensitivity))); (((“Large Vessel Occlusion”) AND (“artificial intelligence”) AND (CTA)) AND ((patients) AND (Triage))).

Studies published between 2019 and 2023 examining LVO detection utilizing AI and CT head scans were considered eligible Table 1. By employing the specified search criteria, 1,528 articles were identified. Two independent reviewers, BB and MB, screened the articles to determine eligibility based on the inclusion criteria. Out of the initial pool, thirty-eight papers were deemed eligible for inclusion in the study. In cases of disagreement, a third reviewer, NY, served as a tiebreaker to reach a consensus.

**Table 1.** Studies.

| Study Year | Title | Study Type | Patient Sample Size | Mean Age | Gender | Ethnicity | Stroke Center |
| --- | --- | --- | --- | --- | --- | --- | --- |
| (Morey, 2020) | Impact of Viz LVO on Time-to-Treatment and Clinical Outcomes in Large Vessel Occlusion Stroke Patients Presenting to Primary Stroke Centers | RCT | 42 | 74 | Female Pre 45% (9/20)<br>Post 50% (11/22; p-value 0.7675 | N/A | Comprehensive |
| (Hassan AE, 2023) | The implementation of artificial Intelligence significantly reduces door-in-door-out times in a primary care center prior to transfer | RCT | 63 | 69 | Female 55.56% | N/A | Single Spoke Primary Care Center |
| (Matsoukas, 2022) | AI Software Detection of Large Vessel Occlusion Stroke on CVA: Real World Diagnostic Test Accuracy Study | Cluster Randomized trial | 1774 | 72 | Females N=877 (49.4%)<br>Females LVO 75 (52.8%) | N/A | Multi-Center |
| (AE, 2020) | Early experience utilizing artificial Intelligence shows significant reduction in transfer times and length of stay in a hub and spoke model | Retrospective Multicenter | 43 | 72 | Female 51.16%<br>Male 48.84% | 23.26% White; 76.74% Hispanic; 0% African American; 0% Asian | Hub and Spoke |
| (Dornbos, 2020) | Automated large vessel occlusion by artificial Intelligence improves stroke workflow metrics: 1st 100 patient experience in a hub and spoke stroke system | Retrospective | 680 | 69 |  | N/A | Comprehensive |
| (JR, 2021) | Real-World Experience with Artificial Intelligence-Based Triage in Transferred Large Vessel Occlusion Stroke Patients | Retrospective | 55 |  | Male 50.0% (13/26) | N/A | Comprehensive |
| (Elijovich L, 2022) | Automated emergent large vessel occlusion detection by artificial Intelligence improves stroke workflow in a hub and spoke stroke system of care | Retrospective | 680 | 70 | Female 64% AI = 29/45<br>Females 54% Non-AI = 32/59 | Non-white race, n (%) AI = 14/43 (32.6) / usual care = 25/59 (42.4) 0.314 | Comprehensive Stroke Center<br>Hub and Spoke |
| (Matsoukas, 2023) | Artificial Intelligence-Assisted Software Significantly Decreases All Workflow Metrics for Large Vessel Occlusion Transfer Patients within a Large Spoke and Hub System | Retrospective | 78 | N/A | Female 53.8% | N/A | Hub and Spoke |
| (Hassan, 2022) | Artificial Intelligence-Parallel Stroke Workflow Tool Improves Reperfusion | Retrospective | 188 | 69 | Female 42.0% | Pre-AI -- Post-AI White 16 (18.6%) -- 26 | Hub and Spoke |
|  | Rates and Door-In to Puncture Interval |  |  |  |  | (25.5%)<br>Hispanic 68 (79.1%)-- 78 (76.5%)<br>Black 1 (1.2%)-- 0 (0.0%)<br>Asian 1 (1.2%)-- 0 (0.0%) |  |
| (Sevilis, 2023 ) | Validation Of Artificial Intelligence to Limit Delays in Acute Stroke Treatment and Endovascular Therapy (VALIDATE) | Retrospective | 14116 | N/A | N/A | N/A | Multicenter (166 Centers total) |
| (Al-Kawaz, 2021 ) | Impact of RapidAI mobile application on treatment times in patients with large vessel occlusion | Retrospective | 64 | N/A | Female: 46.9% non-AI 15; 51.6% AI 16 | N/A | Comprehensive |
| (G, 2022) | Automated Large Artery Occlusion Detection in Stroke: A Single-Center Validation Study of an Artificial Intelligence Algorithm | Retrospective | 595 | N/A | Male 51.4%, N = 306 | N/A | Single Center |
| (A, 2021) | Evaluation of Artificial Intelligence-Powered Identification of Large-Vessel Occlusions in a Comprehensive Stroke Center | Retrospective | 1167 | 62 | Male 59% | N/A | Comprehensive stroke center |
| (M, 2020 ) | Deep Learning Based Software to Identify Large Vessel Occlusion on Noncontrast Computed Tomography | Retrospective | 1453 | 71 | Female 52.4% | N/A | Comprehensive Stroke Centers |
| (BACM, 2022 ) | Artificial intelligence software for diagnosing intracranial arterial occlusion in patients with acute ischemic stroke | Retrospective | 474 | 72 | 235 Males | N/A | Primary Stroke Center |
| (M, 2023) | Real-World Performance of Large Vessel Occlusion Artificial Intelligence-Based Computer-Aided Triage and Notification Algorithms-What the Stroke Team Needs to Know | Retrospective | 618 at Hospital A<br>618 at Hospital B | 74 Hospital A<br>68 Hospital B | Female 43.5% at Hospital A<br>Female 50.2% at Hospital B | Hospital A: Asian (4%), Black or African American (3.1%), white or Caucasian (88.3%), other (3.1%)<br>Hospital B: Asian (5.5%), Black or African American (9.4%), white (69.7%), other (15.4%) | Two Separate Stroke Centers |
| (Jacob, 2022) | Head-to-head comparison of commercial artificial intelligence solutions for detection of large vessel occlusion at a comprehensive stroke center | Retrospective | 263 | 68 | 122 Females | N/A | Single Center, Comprehensive Stroke Center |
| (Shalitin, 2020 ) | AI-powered stroke triage system performance in the wild | Retrospective | 2,544 | 66 | Female 52.48%<br>Male 46.62% | N/A | Multiple Hospital Systems 139 sites [PSC = 73 sites; CSC = 34 sites; Other or unspecified = 32 sites] Sites with CTP available 45.32% (63/139) |
| (JE, 2023 ) | Impact of an automated large vessel occlusion detection tool on clinical workflow and patient outcomes | Retrospective | 439 cases in the pre-AI group<br>321 cases in the post-AI group | N/A | N/A | N/A | Comprehensive Stroke Center |
| (Hassan, 2021 ) | Implementation of Artificial Intelligence Software Significantly Improves Door-In to Groin Puncture Time Interval and Recanalization Rates | Retrospective | 188 | 69 | Female 42.0% | White – Pre-AI 16(18.6%)/post-AI 26 (25.5%)<br>Hispanic – Pre-AI 68 (79.1%) / Post-AI 78 (76.5%)<br>African American – Pre-AI 1 (1.2%)/post-AI 0%<br>Asian Pre-AI 1 (1.2%)/post-AI 0% | Comprehensive Care Center |
| (Wolfe, 2023 ) | Practical performance of stroke detection by Rapid and Viz.AI artificial intelligence applications. | Retrospective | 672 | N/A | N/A | NA | N/A |
| (Devlin, 2019 ) | Automated Detection, Identification, Selection, and Triage using Artificial Intelligence in Large Vessel Occlusions Requiring Critical and Timely Intervention | Multicenter Observational trial | 21 | N/A | N/A | N/A | N/A |
| (Nagamine, 2022) | Evaluation Of the Implementation of An Ai Tool for Large Vessel Occlusion: Impact on Radiologists' Workflow And Patient Outcomes | Retrospective | 144 | N/A | N/A | N/A | N/A |
| (KG, 2021) | Cost-effectiveness of artificial intelligence aided vessel occlusion detection in acute stroke: an early health technology assessment. | Retrospective | 71840 | 66 | N/A | N/A | UK Stroke Registry Data |
| (Sawicki, 2021 ) | Diagnostic Value of Artificial Intelligence—Based Software in Detection of Large Vessel Occlusion in Acute Ischemic Stroke | Retrospective | 108 | 70 | Females 50.9% | N/A | N/A |
| (SE, 2023 ) | Duration and accuracy of automated stroke CT workflow with AI-supported intracranial large vessel occlusion detection. | Retrospective | 100 | 64 | Male 57% | N/A | N/A |
| (M, 2022 ) | Utilization of Artificial Intelligence-based Intracranial Hemorrhage Detection on Emergent Noncontrast CT Images in Clinical Workflow. | Retrospective | 3017 | 66 | Female 47.5% | N/A | Emergency Department |
| (P, 2022) | Validation of a machine learning software tool for automated large vessel occlusion detection in patients with suspected acute stroke. | Retrospective | 217 | N/A | Female 97 (44.7%) | N/A | N/A |
| (T, 2022) | Validation of an artificial intelligence solution for acute Triage and rule-out normal of non-contrast CT head scans | Retrospective | 390 | 54 | N/A | N/A | N/A |
| (RA, 2021 ) | Validation of an artificial intelligence-driven large vessel occlusion detection algorithm for acute ischemic stroke patients. | Retrospective | 202 | 70 | Male 48.5% | N/A | Comprehensive Stroke Centre |
| (A, 2023) | Viz LVO versus Rapid LVO in detection of large vessel occlusion on CT angiography for acute stroke | Retrospective | 360 | N/A | N/A | N/A | N/A |
| (ME, 2023) | Viz.ai Implementation of Stroke Augmented Intelligence and Communications Platform to Improve Indicators and Outcomes for a Comprehensive Stroke Center and Network | Retrospective | 82 | N/A | N/A | N/A | Comprehensive Stroke Centre |
| (K, 2023) | (VISION-S): Viz.ai Implementation of Stroke augmented Intelligence and communications platform to improve Indicators and Outcomes for a comprehensive stroke center and Network - Sustainability | Retrospective | 150 | N/A | N/A | N/A | CSC Centers |
| (Field, 2023) | Artificial Intelligence improves transfer times and ischemic stroke workflow metrics | Retrospective | 262 | N/A | Male 58.8% | N/A | Regional CSC |
| (Jabal, 2023) | Automated CT angiography collateral scoring in anterior large vessel occlusion stroke: A multireader study | Retrospective | 56 | 66 | Female 53% | N/A | Institutional - multireader study |
| (Bruggeman, 2022) | Automated Detection and Location Specification of Large Vessel Occlusion on Computed Tomography Angiography in Acute Ischemic Stroke | Retrospective | 364 | 67 | Male N=185 | N/A | Comprehensive Stroke Center |
| (Martinez - Gutierrez, 2023) | Automated Large Vessel Occlusion Detection Software and Thrombectomy Treatment Times: A Cluster Randomized Clinical Trial | Stepped wedge cluster randomized trial (SW-CRT) | 243 | N/A | Female 50%<br>N=122 | African American 69 (28%)<br>Asian 13 (5%)<br>Hispanic 41 (17%)<br>White 108 (44%)<br>Other 12 (5%) | Comprehensive Stroke Centers |

Legend: RCT: Randomized Control Trials, PSC: Primary Stroke Center, CSC: Comprehensive Stroke Center

Inclusion criteria

- Stroke, large vessel occlusion
- CT/angiogram/perfusion
- Artificial Intelligence
- Deep and machine learning
- AI model performance metrics and costs
- Patient processing times
- Operations Management

**Exclusion Criteria:**

- Patients presenting with intracranial hemorrhages
- Patients with aneurysms
- MRI studies
- Pre-hospital AI triaging systems studies
- Mobile stroke unit studies
- Pediatric patients
- Review Articles

Grammarly version 1.2.63.1310 and Notion AI were utilized for manuscript editing purposes.

Meta-analysis was conducted utilizing a random effects model, and the I² statistic was applied to assess the heterogeneity among individual studies. To evaluate potential publication bias, funnel plots were generated. The analysis was carried out using the “Comprehensive Meta Analysis v. 4 “software.

## Results 3.0

This study compared AI-augmented LVO detection and non-AI in various patient processing times for emergent endovascular therapy in acute ischemic strokes. The results indicated significant differences between the two groups across several parameters.

The utilization of AI demonstrated higher sensitivity in detecting stroke compared to non-AI methods with an OR of 0.91 (95% CI: 0.88 - 0.95, p < 0.001), suggesting improved sensitivity in AI-augmented LVO detection; an NIHSS score was 16.20 (95% CI: 14.96 - 17.45, p = 0.001), indicating the severity of neurological impairment in the study population.

Triage Time, Door-to-INR, and Door-to-Arterial Puncture Time all showed statistically significant odds ratios (OR) in favor of AI utilization, with values of 0.39 (95% CI: 0.29 - 0.54, p < 0.001), 0.30 (95% CI: 0.21 - 0.42, p < 0.001), and 0.50 (95% CI: 0.30 - 0.82, p = 0.007), respectively. These findings suggest that using AI was associated with reduced times in these domains, translating to more rapid diagnosis and treatment.

Similarly, the CT-to-Puncture Time was quicker when AI was utilized than in cases without AI intervention, with an odds ratio (OR) of 0.57 (95% CI: 0.31-1.04, p = 0.065). However, this association did not achieve statistical significance.

The Last Known Well (LKW) to Time of Arrival resulted in an OR of 1.15 (95% CI: 0.83-1.59, p = 0.409). Meanwhile, the Door-to-CTA Time yielded an odds ratio (OR) of 0.77 (95% CI: 0.37-1.60, p = 0.489). However, neither association reached statistical significance. Patient Transfer Times between primary and comprehensive stroke centers did not differ significantly between AI and non-AI conditions, with an OR of 0.98 (95% CI: 0.73 - 1.32 p = 0.893). Similarly, DIDO Time did not show a significant association with AI utilization, yielding an OR of 1.19 (95% CI: 0.21 - 6.88, p = 0.848).

The studies included in this meta-analysis did not provide insights into cost-effectiveness or mortality.

## Discussion 4.0

Implementing AI for detecting LVO in time-sensitive scenarios has offered valuable insights into its effectiveness in acute stroke care management. Our findings have significant implications, such as AI-driven, timely stroke detection and intervention, which can be crucial in optimizing outcomes in stroke cases. A notable difference was observed between AI and traditional non-AI methods across several critical patient processing times.

For instance, Triage Time, a pivotal component of initial patient assessment, exhibited a significantly reduced odds ratio (OR) of 0.39 (95% CI: 0.29 - 0.54, p < 0.001) with AI integration, indicative of expedited evaluations. Similarly, the Door-to-INR Notification Time reflected an OR of 0.30 (95% CI: 0.21 - 0.42, p < 0.001), suggesting quicker notification of the endovascular team, which could lead to timelier endovascular therapy. CT-to-Arterial-Puncture Time (OR = 0.57, 95% CI 0.31 - 1.04, p = 0.065) exhibited considerable variability in its impact across populations, with a wide interval (0.05 to 6.14) suggesting diverse outcomes influenced by AI. Hospital-specific factors could have influenced this outcome. While the Door-to-Arterial Puncture Time favored AI with an OR of 0.50 (95% CI: 0.30 - 0.82, p = 0.007), the 95% CI range (0.09 to 2.89) signifies potential variability within patient cohorts. It is necessary to identify hospital-specific factors that could influence this outcome.

The reduction in Triage Time, Door-to-INR Notification Time, CT-to-Arterial Puncture, and Door-to-Arterial Puncture Time in the AI groups implies the potential for AI interventions to enhance stroke team responsiveness, thereby potentially minimizing adverse clinical outcomes and optimizing the treatment window for stroke care.

Conversely, the patient’s LKW to Time-of-Arrival OR of 1.15 (95% CI: 0.83 - 1.59, p = 0.409) and DIDO Time OR of 1.19 (95% CI: 0.21 - 6.88, p = 0.848) and inter-facility Transfer Times between primary and comprehensive stroke centers an OR of 0.98 (95% CI: 0.73 - 1.32, p = 0.893), demonstrated a relatively marginal and statistically insignificant discrepancy between AI and non-AI methodologies. Similarly, Door to CTA Time, with an OR of 0.77 (95% CI: 0.37 - 1.60, p = 0.489), displayed a wide range in 95% of comparable populations (0.00 to 4419.82). There seems to be no apparent explanation for AI to reduce these times, given that AI is typically initiated after the completion of the CT scan. However, hospitals equipped with AI may operate more efficiently due to enhanced resources, potentially leading to quicker patient triage and imaging processes. These fluctuations indicate the multifaceted nature of stroke management, prompting further investigation into unexplored factors and confounding variables contributing to temporal disparities.

There are key factors contributing to delays in acute stroke care during the transfer from a primary stroke center to a comprehensive stroke center, including: 1) Distance: Geographical distance between primary and comprehensive stroke centers can significantly impact transfer times, especially in rural areas with limited access to healthcare facilities (Ifejika, 2021). 2) Transportation logistics: Availability and coordination of transportation, such as ambulances or air transport, can influence transfer efficiency. Delays may occur due to logistical challenges, including traffic congestion or adverse weather conditions, time of day, and visibility (Glober, 2023). 3) Interfacility communication: Effective communication between the primary and comprehensive stroke centers is crucial for seamless transfer and continuity of care (Patterson, 2023). Delays may arise from communication barriers or breakdowns in information exchange between healthcare providers (Patterson, 2023). 4) Bed availability: Limited beds or resources at the comprehensive stroke center may lead to delays in accepting transferred patients. Capacity constraints can exacerbate delays, particularly during peak demand periods (Monks, 2016). Transferring the patient to the accepting facility’s emergency room helps mitigate this issue, as the patient does not require a traditional hospital bed but can proceed directly to the angiography suite. Following the procedure, they can be moved to a post-operative area until an ICU bed becomes available. 5) Transfer protocols: Adherence to standardized protocols and guidelines is essential for expediting the transfer process (Kodankandath, 2017). Delays may occur if transfer protocols are not followed or if there are discrepancies in protocols between facilities (Kodankandath, 2017). 6) Specialist availability: Timely access to stroke specialists, neurologists, and other healthcare professionals is critical for prompt evaluation and treatment at the comprehensive stroke center (Patterson, 2023). Delays may arise if specialist expertise is unavailable or there are consultation delays (Patterson, 2023). This issue can be effectively addressed through dedicated telestroke services. Telestroke services offer expedited stroke detection and treatment. 7) Diagnostic imaging: Availability and interpretation of diagnostic imaging studies, such as computed tomography (CT) scans or magnetic resonance imaging (MRI), are essential for accurate stroke diagnosis and treatment planning. Delays may occur if there are technical issues with imaging equipment or delays in image interpretation (Yu, 2002). 8) Treatment protocols: Consistency in treatment protocols between primary and comprehensive stroke centers is vital for ensuring timely and appropriate interventions. Delays may arise if there are differences in treatment protocols or delays in implementing treatment plans (Zackrison, 2019). 9) Patient condition: The clinical stability of the patient and the urgency of treatment can impact transfer prioritization and timing. Delays may occur if there are complications or changes in the patient’s condition during transfer, requiring additional stabilization or interventions (Ali, 2018). 10) Regulatory requirements: Compliance with regulatory requirements, such as transfer agreements or documentation standards, is necessary for legal and regulatory compliance (Higashida, 2013). Delays may occur if there are discrepancies or delays in fulfilling regulatory requirements for patient transfer (Higashida, 2013). Addressing these key factors may improve patient processing times but requires a multidisciplinary team approach involving collaboration between healthcare providers, administrators, and transportation agencies to optimize acute stroke care and minimize delays from primary stroke centers to comprehensive stroke centers (Higashida, 2013).

AI Sensitivity demonstrated an OR of 0.91 (95% CI: 0.88 - 0.95, p < 0.001), affirming the ability of AI systems to identify stroke. The average NIHSS of 2.240 post-AI augmentation (95% CI: 0.943 - 3.536, p = 0.001) underscores the clinical viability of AI algorithms for timely LVO identification. Furthermore, it is essential to acknowledge that there may be variability in performance metrics for AI software in stroke detection across different software companies, emphasizing the need for careful consideration when making clinical decisions (Murray, Volume 12). Sensitivity, specificity, accuracy, speed of analysis, and AI’s communication of findings to all on-call stroke team members are the key measures to consider when evaluating the performance of different software systems.

Most studies in this manuscript comprise a combination of retrospective and prospective study designs. Over time, we expect more prospectively designed clinical trials to emerge. Our results suggest that implementing AI technologies can enhance operational efficiency for hospital systems, with notable improvements observed in reducing the length of stay through AI-augmented decision-making by healthcare providers. Furthermore, these AI-augmented workflows may potentially influence clinical outcomes positively. However, our study did not address the impact of AI implementation on costs and clinical outcomes, primarily due to the nascent nature of AI technology, with outcome and cost studies expected soon. In the near future, we anticipate more prospective randomized controlled trials equipped with robust methodologies to detect AI-augmented influence on clinical outcomes and costs.

## Limitation 5.0

The field of AI is evolving rapidly, and it is possible that not all pertinent studies conducted after the search period range of the study have been incorporated.

**Figure 1.**
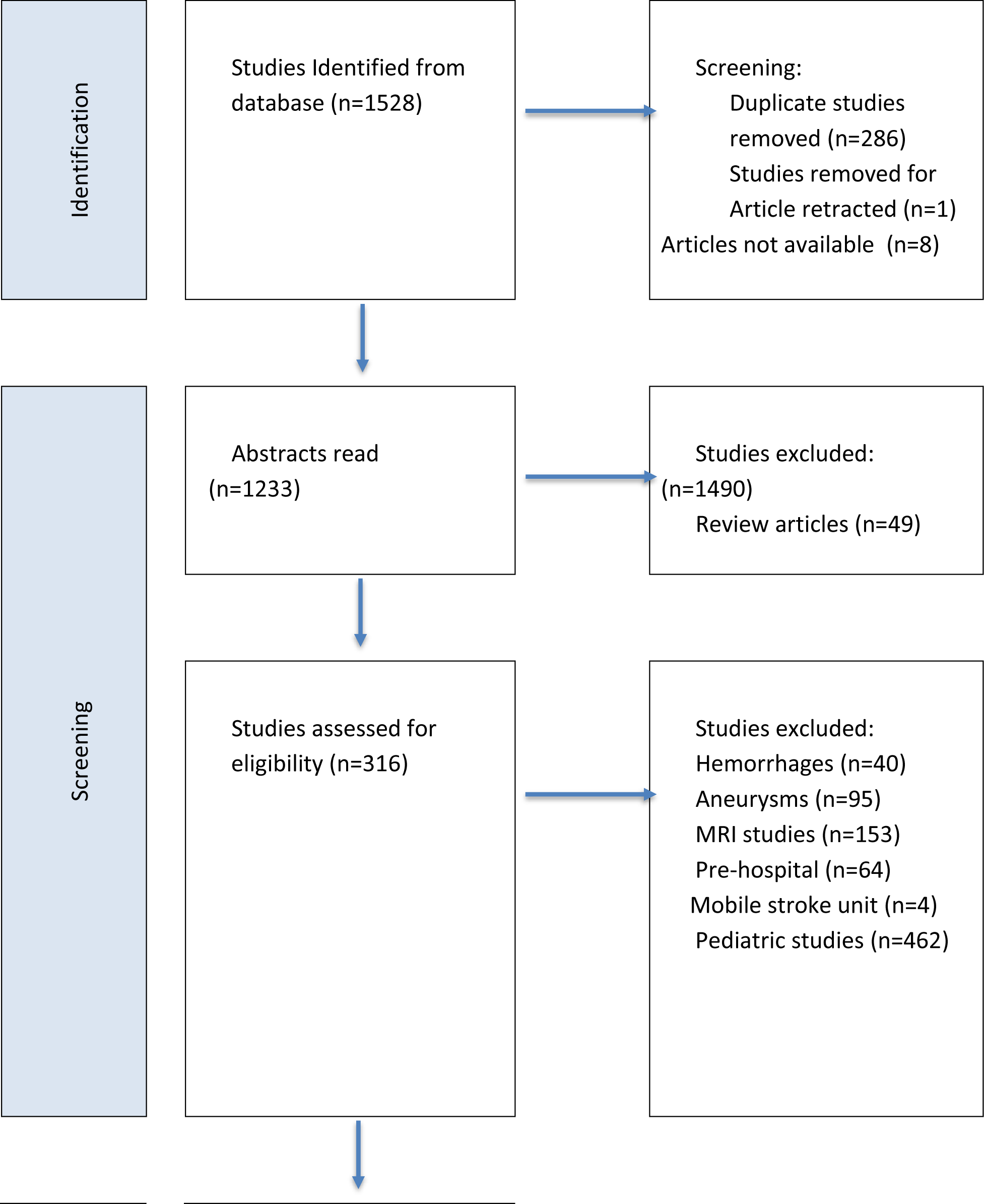
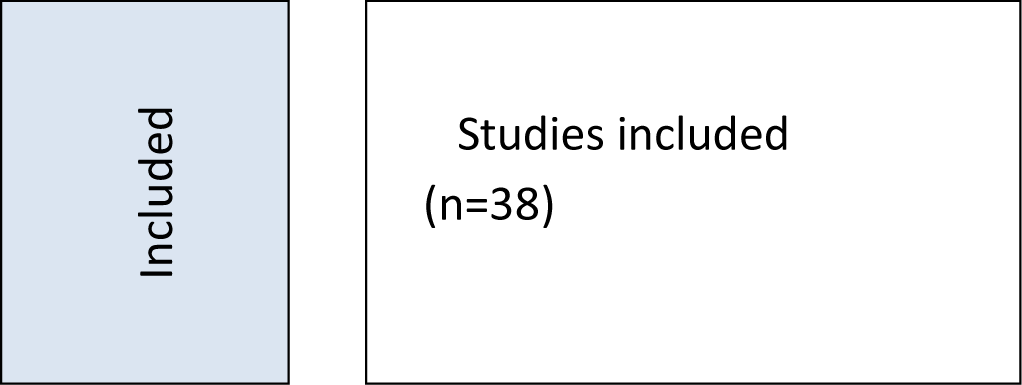
Flow chart of studies using PRISMA guidelines.

**Figure 2.**
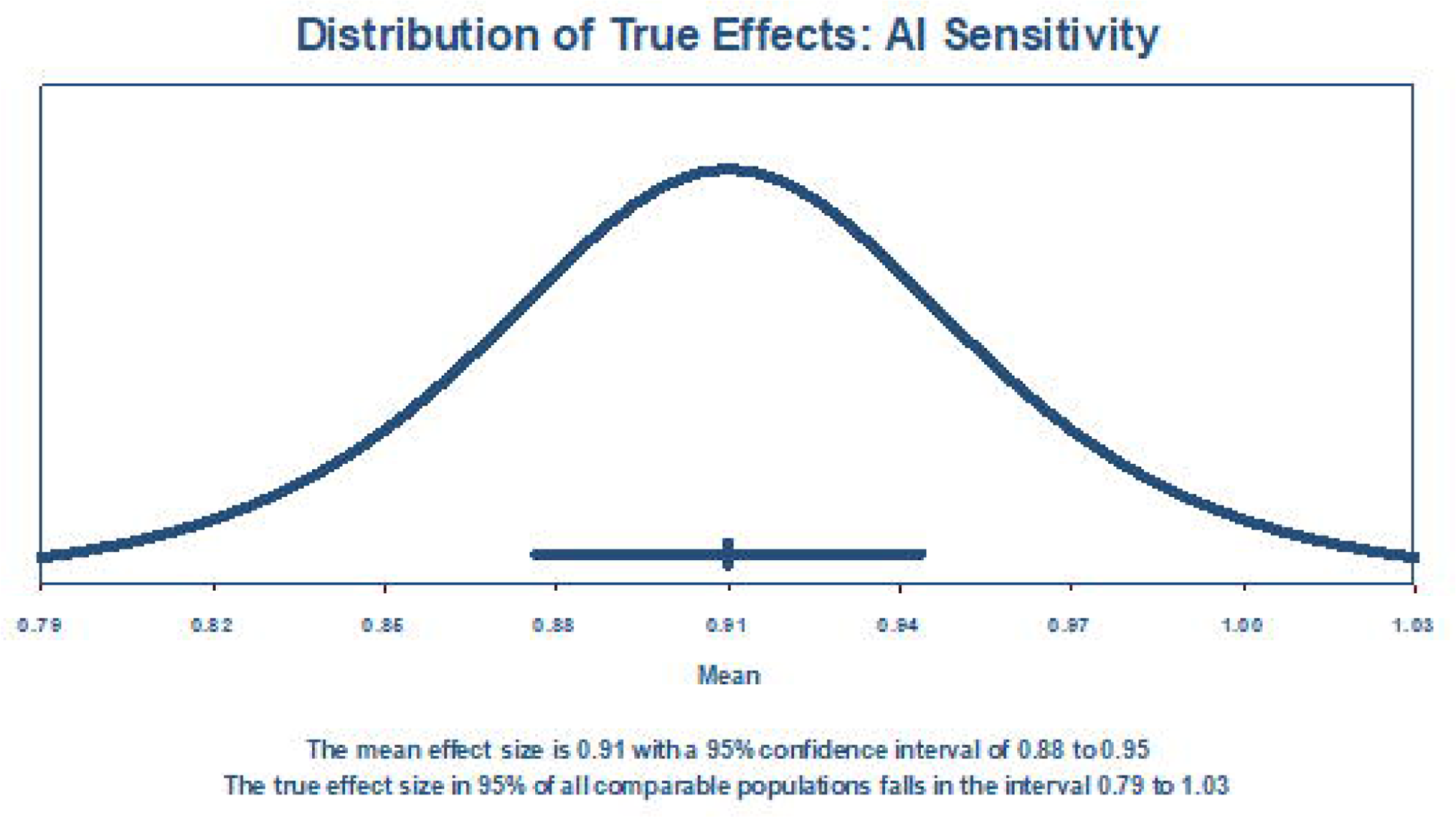
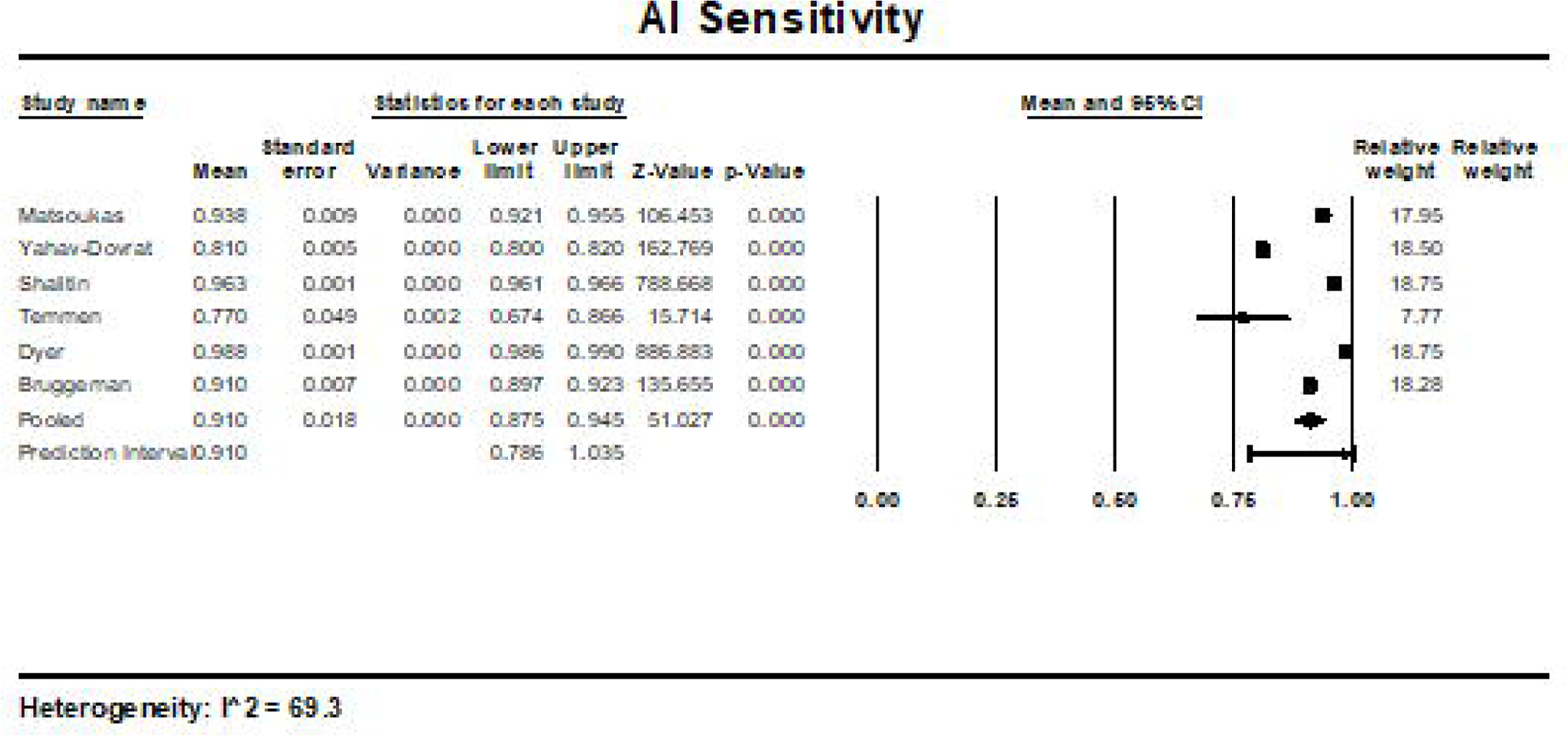
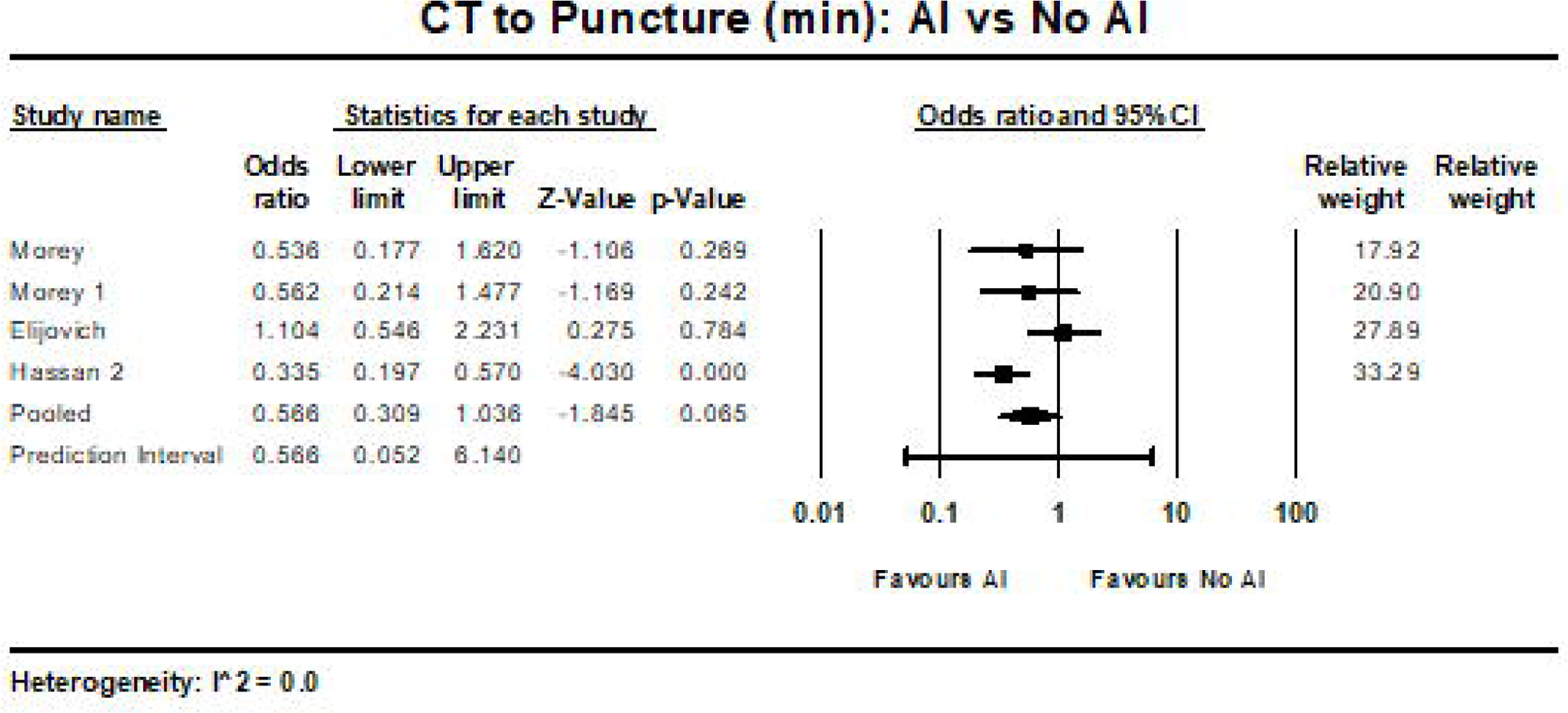
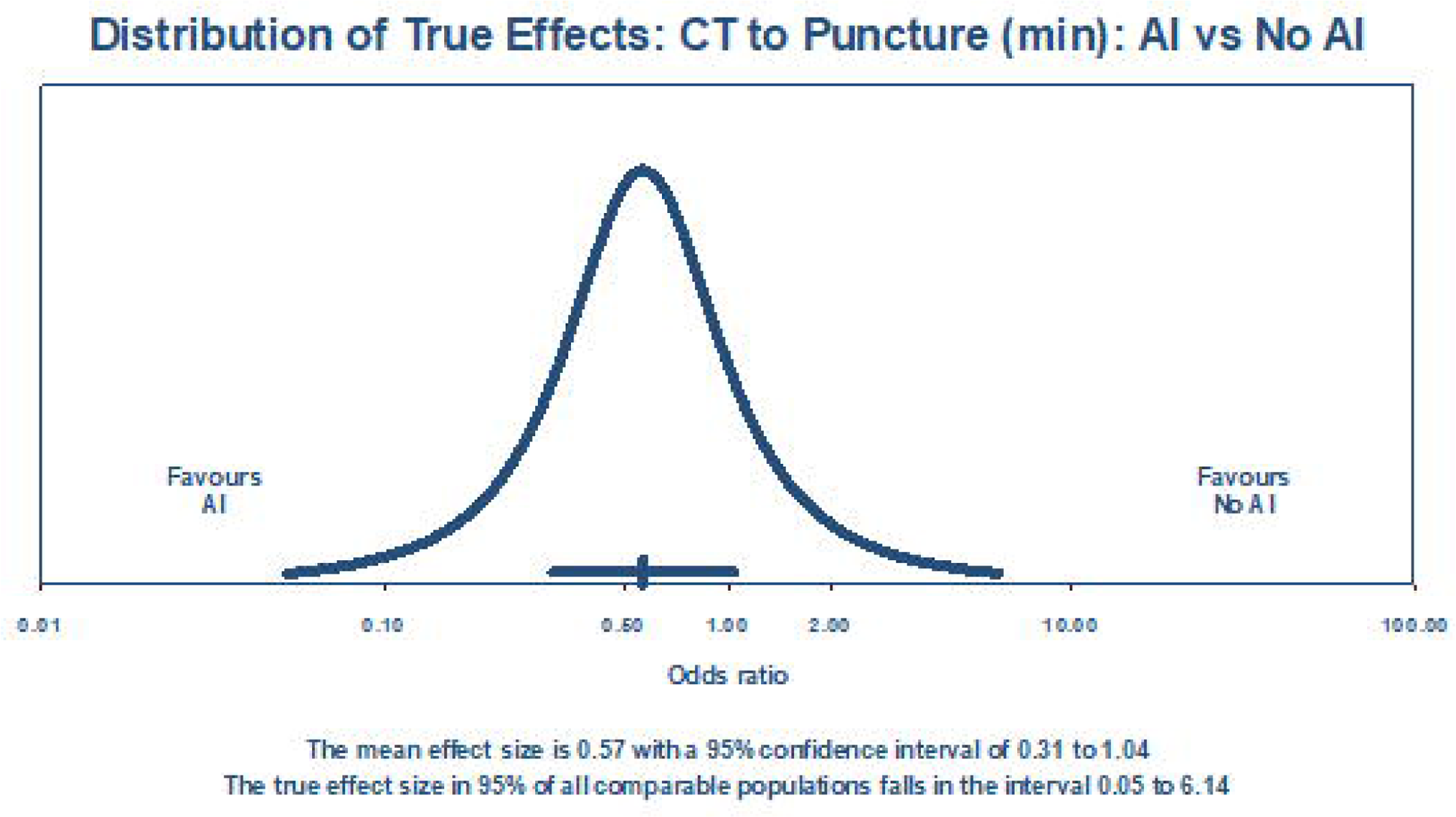
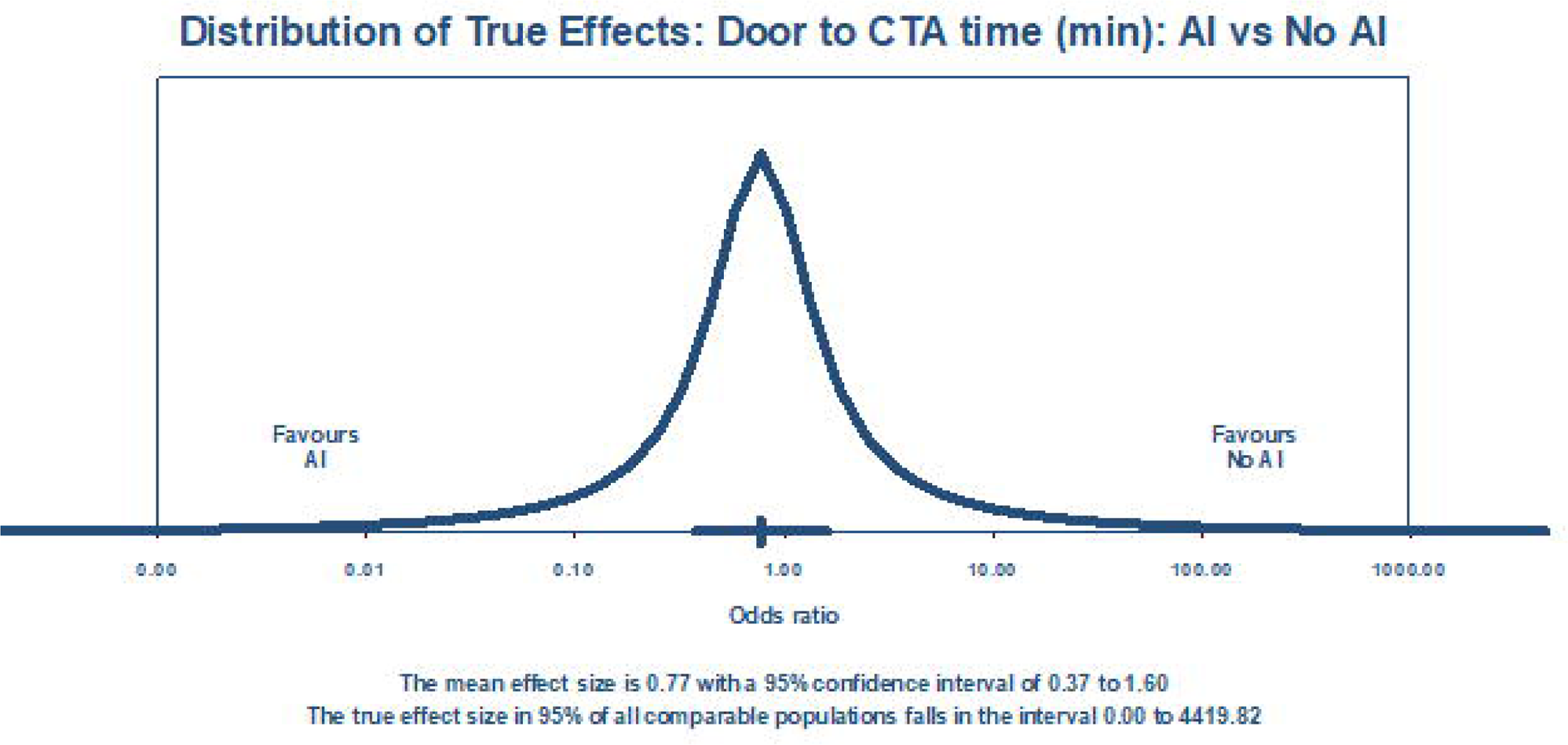
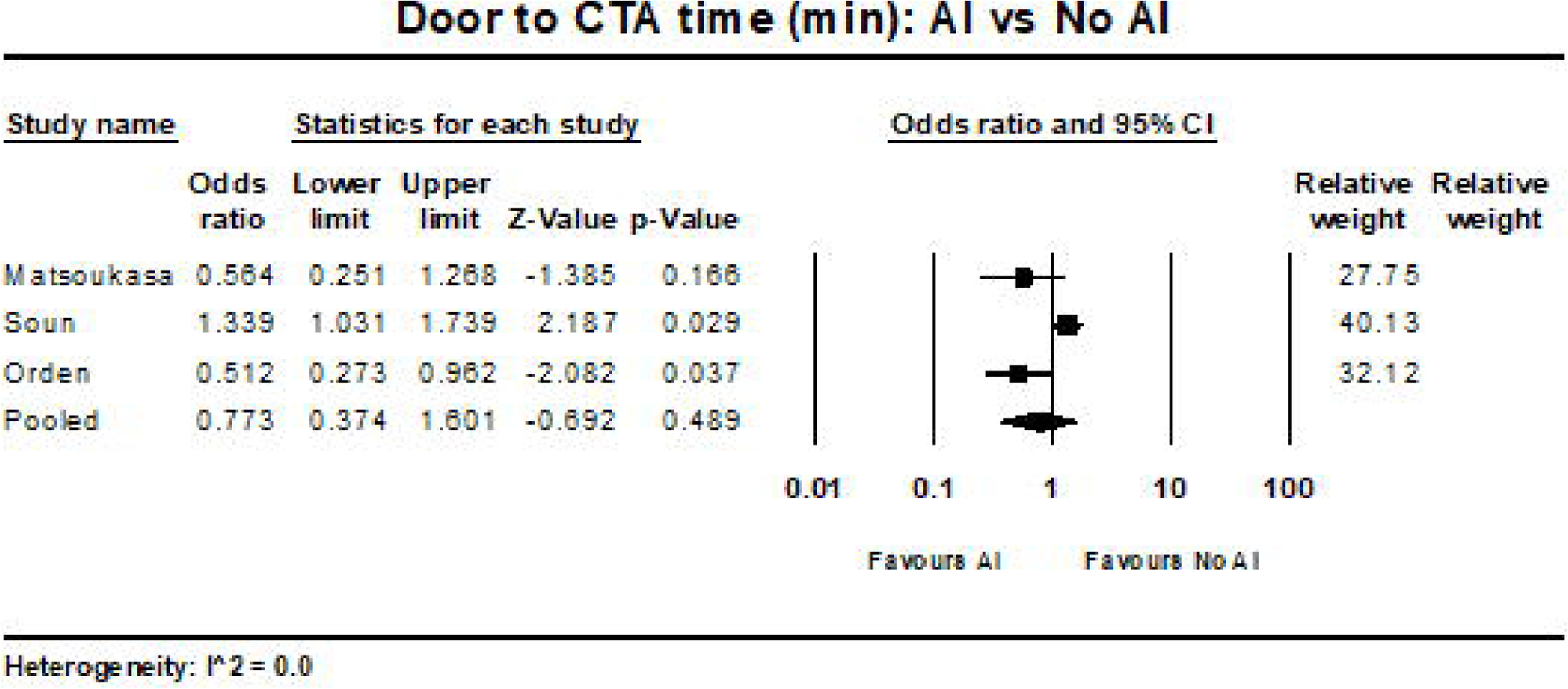
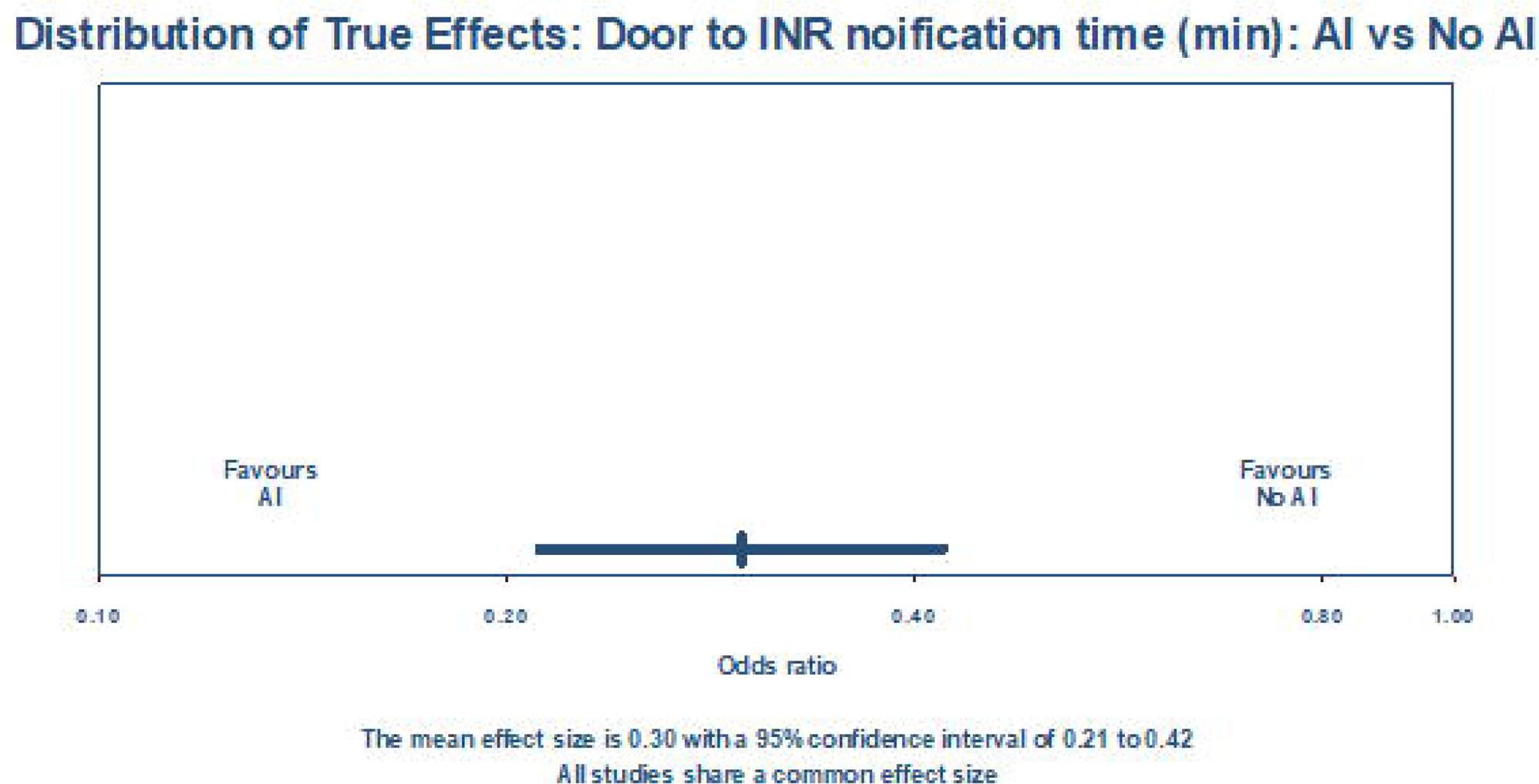
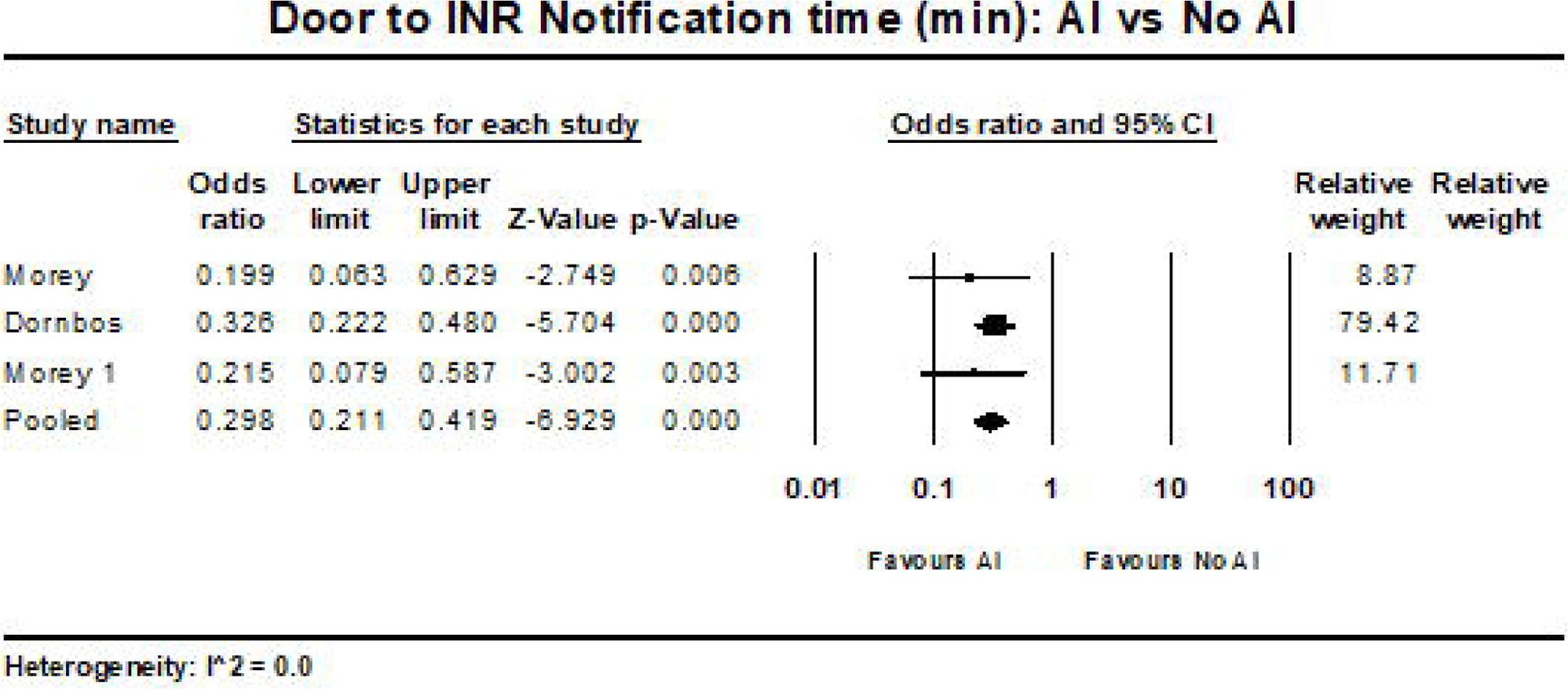
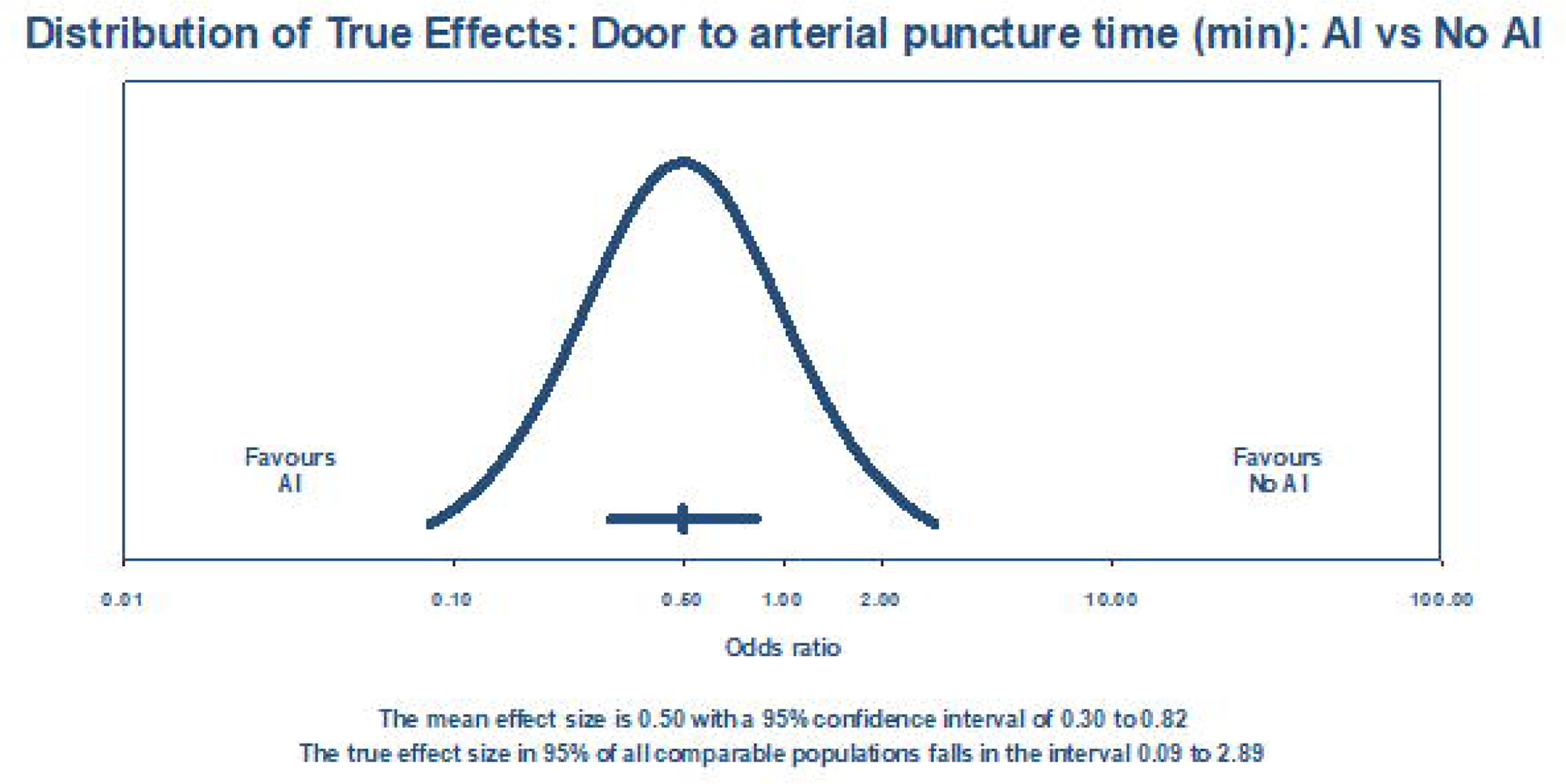
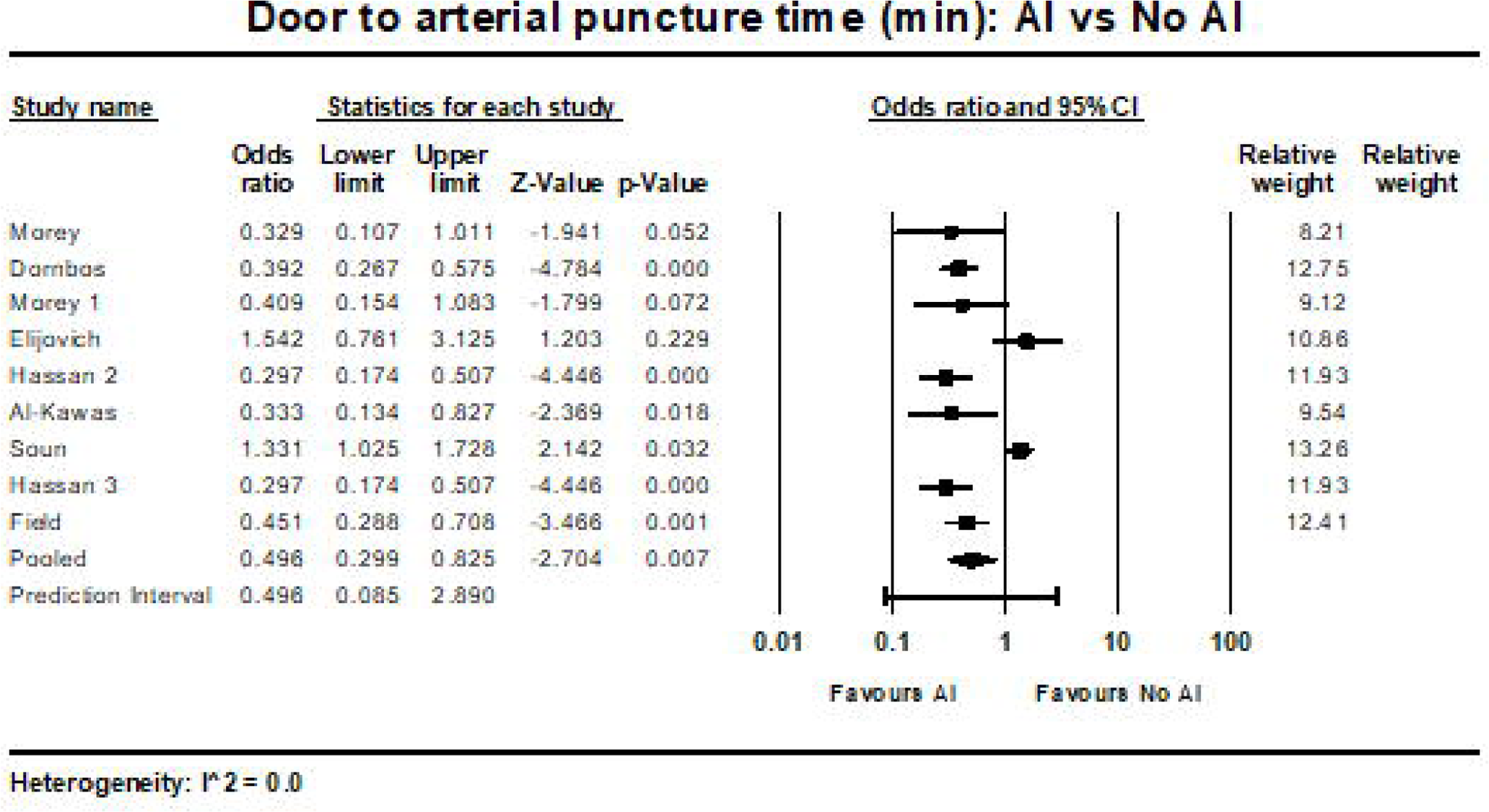
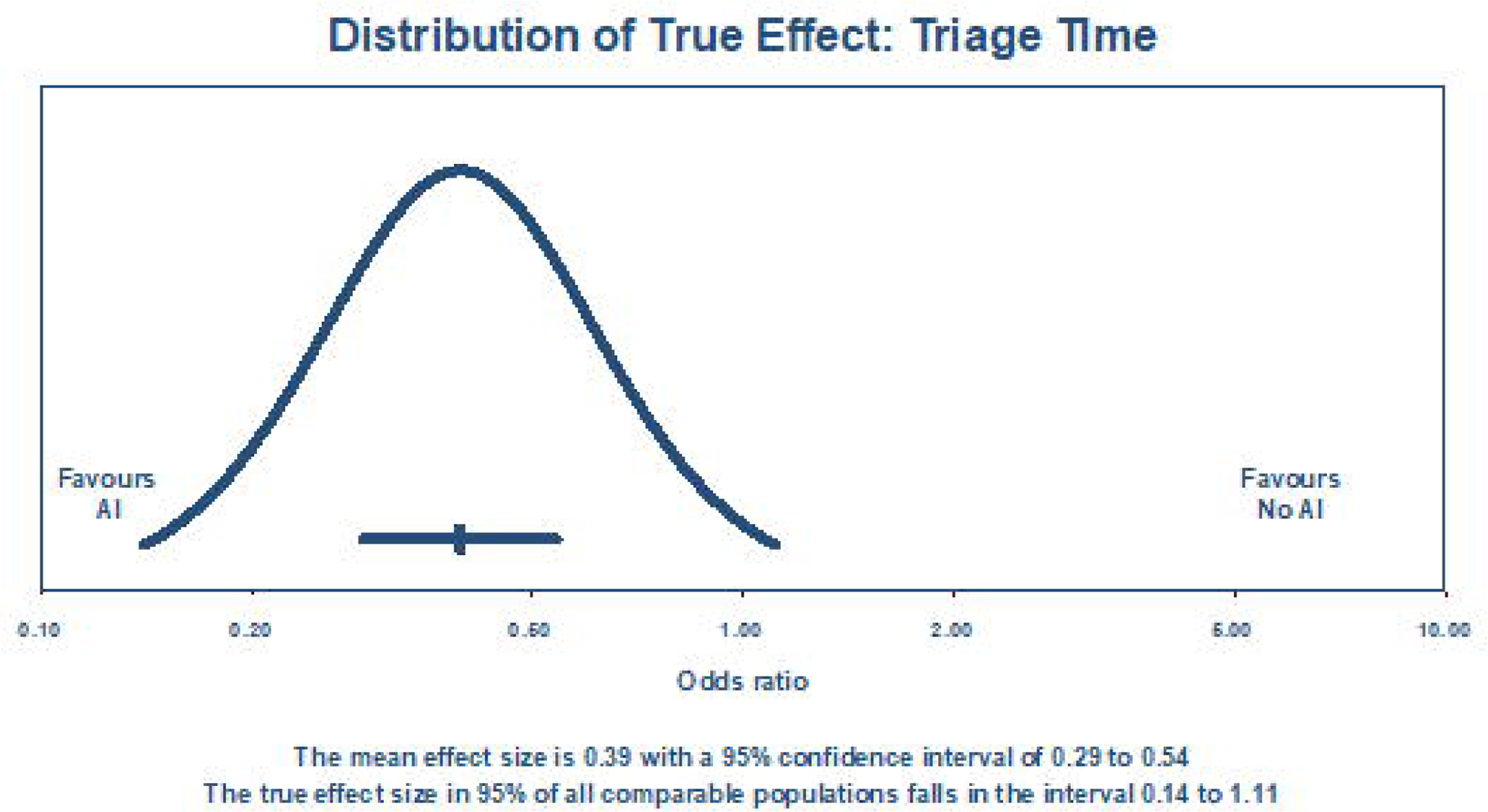
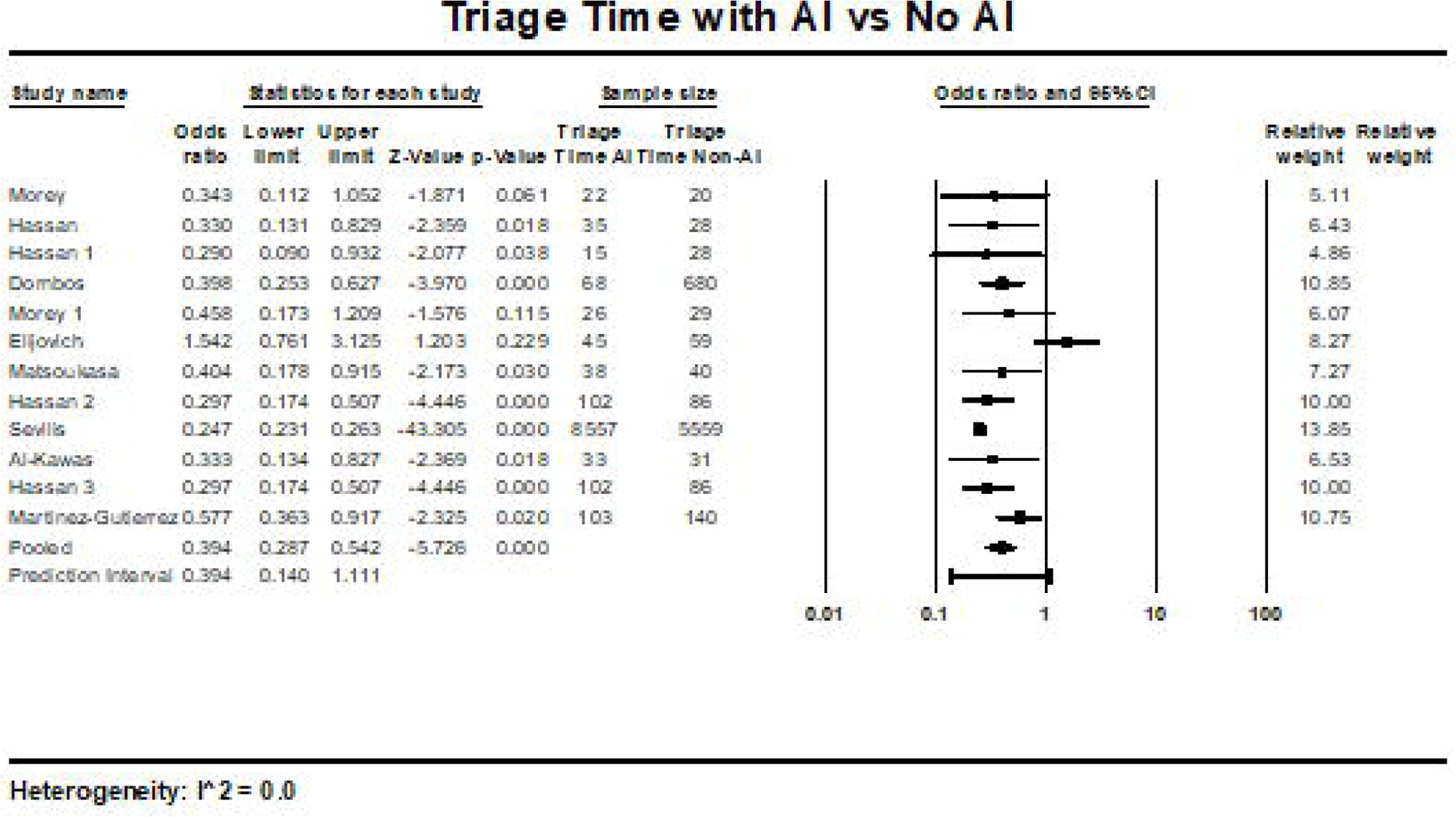

## Data Availability

All data produced in the present study are available upon reasonable request to the authors

## Acknowledgments

Grammarly version 1.2.63.1310 and Notion AI were utilized for manuscript editing purposes.

